# Detecting eczema areas in digital images: an impossible task?

**DOI:** 10.1101/2022.03.03.22271780

**Authors:** Guillem Hurault, Kevin Pan, Ricardo Mokhtari, Bayanne Olabi, Eleanor Earp, Lloyd Steele, Hywel C. Williams, Reiko J. Tanaka

## Abstract

Assessing the severity of atopic dermatitis (AD, or eczema) traditionally relies on a face-to-face assessment by healthcare professionals, and may suffer from inter- and intra-rater variability. With the expanding role of telemedicine, several machine learning algorithms have been proposed to automatically assess AD severity from digital images. Those algorithms usually detect and then delineate (“segment”) AD lesions before assessing lesional severity, and are trained using the data of AD areas detected by healthcare professionals. To evaluate the reliability of such data, we estimated the inter-rater reliability of AD segmentation in digital images.

Four dermatologists independently segmented AD lesions in 80 digital images collected in a published clinical trial. We estimated the inter-rater reliability of the AD segmentation using the intra-class correlation coefficients (ICCs) at the pixel-level and the area-levels for different resolutions of the images. The average ICC was 0.45 (*SE* = 0.04) corresponding to a “poor” agreement between raters, while the degree of agreement for AD segmentation varied from image to image.

The AD segmentation in digital images is highly rater-dependent even between dermatologists. Such limitations need to be taken into consideration when the AD segmentation data are used to train machine learning algorithms that assess eczema severity.

## INTRODUCTION

Atopic dermatitis (AD, also called eczema) is one of the most common chronic skin diseases [1]. Many clinical trials on AD treatment include the assessment of AD severity that changes dynamically over time. The assessment of AD severity usually consists of the visual inspection of eczema lesions by healthcare professionals who grade the intensity of several disease signs (such as dryness, redness, and excoriations) and estimate the area (extent) covered by eczema.

The recent development of machine learning (ML) methods, together with the need for telemedicine, resulted in an increasing interest in developing computer vision algorithms for automatic evaluation of AD severity from digital images [2] [3] [4] [5]. Those algorithms generally consist of two steps: (i) identifying areas covered by eczema in each image - so that the images are “segmented” - either manually as part of the data pre-processing (human-in-the-loop) or automatically by an algorithm, and then (ii) predicting the severity of eczema features in the segmented areas. Therefore, reliable detection of eczema lesions is a prerequisite for assessing the severity of these lesions.

The lack of high-quality segmentation labels is one of the main obstacles to developing machine learning methods in medical applications [6] [7]. If the eczema segmentation data provided by dermatologists are of low quality (noisy labels), the algorithms for automatic detection of AD lesions trained with such data may learn the biases contained in the data. For example, models trained with noisy labels to segment brain lesions required an order of magnitude more data than those trained with accurate labels to achieve similar segmentation performance [7]. Inaccurate eczema segmentation can also have effects on assessing severity in eczema images, as the segmentation may only include severe lesions or specific disease signs. If the areas of dryness are never segmented, for example, the assessments of dryness from the segmented eczema images are likely to be inaccurate. The classification accuracy of cancerous prostate tissue images by ML algorithms was found to be decreased by 10% when trained with data that contained incorrectly labelled images [7].

However, it is still unclear whether high-quality eczema segmentation data can be obtained from dermatologists consistently to the best of our knowledge. Trying to measure the eczema area accurately is challenging in real life due to the ill-defined nature of AD. Charman et al. [8] showed a very poor inter-rater reliability (IRR) with a kappa statistic of 0.09 for in-person scoring of the extent of eczema by six experts on six patients. IRR refers to the degree of agreement between raters, i.e. to what extent the labels (the extent of eczema in the case of [8]) are independent of a particular rater. If IRR is high, “raters can be used interchangeably without the researcher having to worry about the categorization being affected by a significant rater factor” [9]. This study quantified the IRR of eczema area segmentation by four dermatologists in 80 digital images of varying quality from paediatric AD patients collected in a published clinical study [10].

## METHODS

### Data

Our data originate from the SWET trial [10], a randomised controlled trial that investigated the effects of the use of ion-exchange water softeners on the control of AD symptoms. The trial included 310 children aged from 6 months to 16 years with moderate to severe AD, from whom digital images of representative AD sites were taken and the Six Area, Six Sign Atopic Dermatitis (SASSAD) severity score [11] was assessed at multiple clinical visits. The images can be considered “realistic” as they were taken using different devices, contain significant areas of background and vary in resolution and subjective quality, such as focus, lighting and blur. Each image was annotated with the intensity scores (0-3) of the six signs for SASSAD (erythema, exudation, excoriation, dryness, cracking and lichenification) at the corresponding representative AD site. We represented the severity score for each image by the sum of the six intensity scores.

From a total of 1345 eczema images available, we used a random number generator computer program to select 80 images at random without replacement (Table 1). We asked four dermatologists (raters) to segment AD lesions within each image (fully crossed design). One rater (HW, rater 1) is a dermatologist with over 30 years of experience assessing eczema. The other three raters (BO, EE, LS) are trainee dermatologists nearing the end of their training who received feedback from HW on eczema area segmentation beforehand to minimise variation. The raters also assessed the image quality (“low”, “normal”, or “high”) for segmenting eczema areas and optionally reported quality issues (“out-of-focus”, “overexposed”, or “other reasons”). We believe that this group of raters with different levels of clinical experience is realistic and represents a “normal practice” for this type of time-consuming task. The segmentation of skin vs background was performed by a non-clinician (RM) as this task did not require clinical expertise.

**Table 1:** Characteristics of the original dataset and the selected images.

|  | SWET dataset | Selected images |
| --- | --- | --- |
| **Number of patients with images (% female)** | 287 (43%) | 71 (44%) |
| **Patients of (declared) “white” ethnicity (%)** | 223 (78%) | 54 (76%) |
| **Mean age in years (SD)** | 5.6 (4.1) | 5.1 (4.0) |
| **Number of images** | 1345 | 80 |
| **Images of legs (%)** | 534 (40%) | 31 (39%) |
| **Images of hands (%)** | 372 (28%) | 19 (24%) |
| **Images of arms (%)** | 190 (14%) | 14 (17%) |
| **Images of feet (%)** | 148 (11%) | 12 (15%) |
| **Images of the head and neck area (%)** | 53 (4%) | 4 (5%) |
| **Images of the trunk or back (%)** | 48 (3%) | 0 (0%) |
| **Mean regional SASSAD (SD, max=18)** | 6.3 (3.2) | 6.3 (3.2) |
| **Mean TISS (SD, max=9)** | 3.0 (1.8) | 3.2 (1.8) |

Labelbox (https://labelbox.com) was used as the image segmentation tool for its ease of use and accessibility. Examples of segmented image masks are shown in Figure 1.

**Figure 1:**
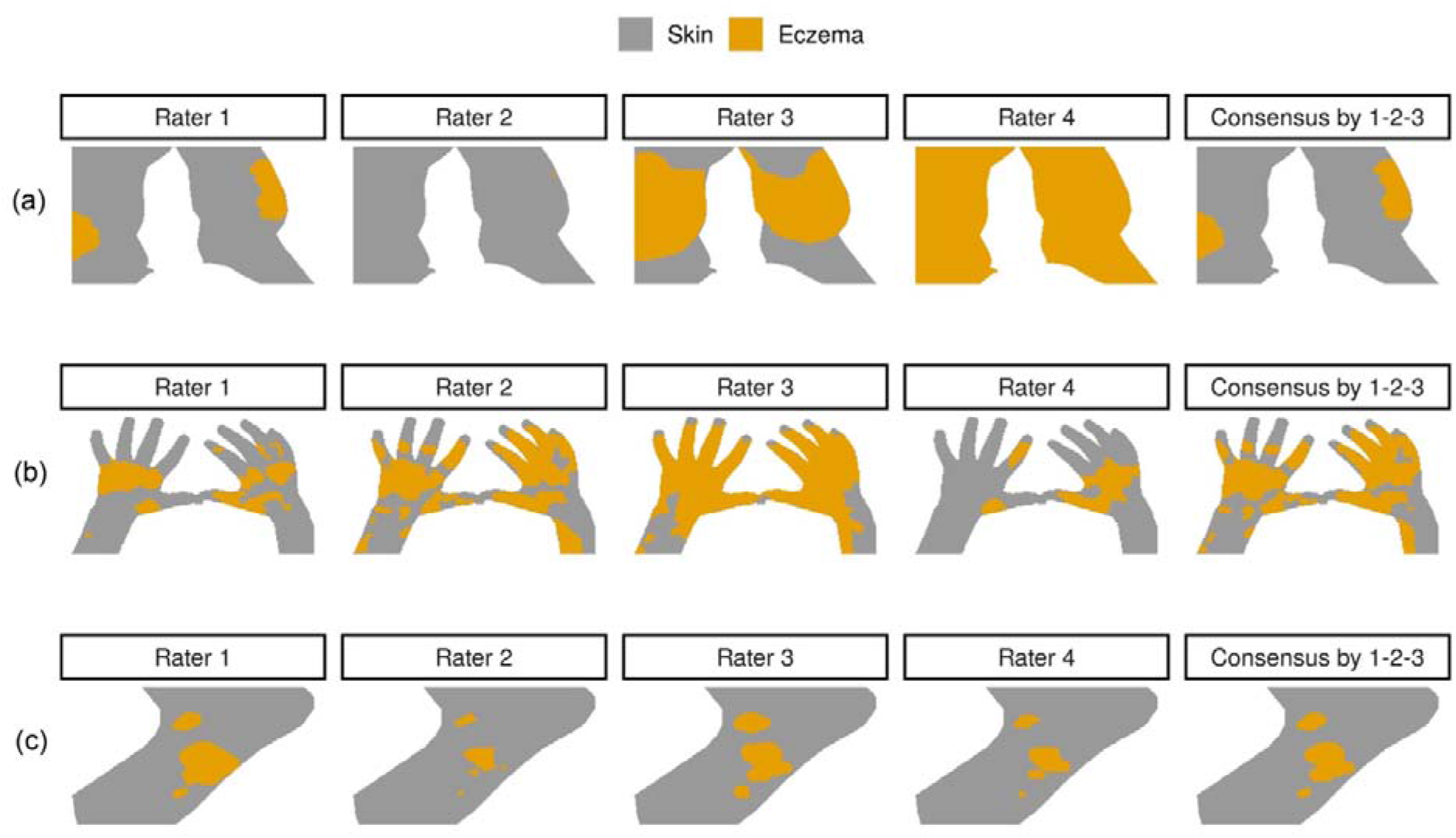
Eczema (orange) and skin (grey) masks for three example images ((a) leg, (b) hands, and (c) foot) segmented by four raters (columns). The last column illustrates a consensus segmentation by the raters 1, 2 and 3. The consensus segmentation was used as a “ground truth” when evaluating the segmentation performance of the rater 4. As an example, the three images represent (a) a very poor IRR (intra-class correlation coefficient (*ICC*) = 0.026, Krippendorff’s alpha (*KA*) = −0.19, (b) an average (i.e. poor in this dataset) IRR (*ICC* = 0.41, *KA* = 0.19), and (c) an excellent IRR (*ICC* = 0.976, *KA* = 0.63).

### Metrics for inter-rater reliability (IRR)

To quantify the IRR, we computed the intra-class correlation coefficient (ICC), which quantifies the association between categorical or continuous ratings assigned to the same rating unit (e.g. the pixel for the pixel-level eczema segmentation). The ICC is defined as the proportion of variance that can be attributed to between-unit variance (e.g. between-pixel variance) to the total variance (e.g. the sum of between-pixel variation and between-rater variance) [12] [13]. It takes values between 0 and 1, with higher ICC values corresponding to better consensus between raters. While the interpretation of ICC values is still debated [14], a consensus is that *ICC* < 0.5 and *ICC* > 0.9 indicate a “poor” and an “excellent” agreement, respectively.

We evaluated the ICC at the pixel- and area-levels for each image. The area-level ICC was calculated to investigate whether our IRR estimates were sensitive to the resolution of the images and whether there was a consensus in identifying larger regions of eczema (or the local extent of eczema) beyond the pixel-level precise segmentation. To estimate the area-level ICC, we compressed the original images to *d* × *d* cells for *d* ∈ {10, 15, 20} and counted the number of eczema pixels in each cell (Figure S1). We also calculated the extent ICC, which is the ICC for the total extent of eczema (proportion of eczema against skin pixels) that can be seen as the limit of the area-level ICC when *d* → 1.

We also reported Krippendorff’s alpha (KA) at the pixel-level. KA is a generalisation of Kappa coefficients (e.g. Cohen’s Kappa, Fleiss’ Kappa) and is suitable for categorical ratings with more than two raters and with a fully crossed design, as in this study [12]. The maximum KA is 1 corresponding to the perfect consensus between raters, and KA=0 corresponds to a chance-level agreement. We did not consider the Intersection over Union (IoU, aka Jaccard index), a similarity metric often used with images, as it does not control for chance-agreement between raters and is limited to two raters.

### Estimating the IRR metrics

In estimating the IRR metrics, we considered only skin pixels so that the chance-agreement does not include background pixels. We analysed 71 images, as we excluded 9 images in which the extent of eczema involvement was more than 95% for 3 or 4 raters, because it implies a high chance-agreement for which an agreement measure cannot be estimated reliably. No images had an extent of eczema less than 5% for 3 or 4 raters.

We investigated whether the IRR estimates were driven by the segmentation of one rater, notably to see whether excluding the most experienced rater (rater 1) influenced the IRR, by conducting a “leave-one-rater-out” sensitivity analysis.

The ICC was estimated using the “rptR” package in R [13]. For both the pixel- and the area-level ICC, we used a mixed-effects logistic regression (binary outcomes of skin/AD) with random effects on raters and pixels/areas (two-way random-effects ICC), and the ICC was calculated on the latent (logit) scale. For the area-level ICC, the model used proportion data as input. For the extent ICC, we computed a two-way random-effects ICC (with an image as the grouping unit) using a mixed-effects linear regression model on the logit of the extent, and confidence intervals were estimated using bootstrap with 1000 resampling. KA was computed using the “irr” package in R.

### Metrics for segmentation performance

To evaluate how much of the segmentation errors can be attributed to the inter-rater reliability, we compared the pixel-level eczema segmentation provided by a specific rater with a “consensus” segmentation and calculated the segmentation performance of the rater compared to their peers. The consensus segmentation was obtained from the other three raters by majority voting, i.e. each pixel was labelled as eczema if at least two out of the three raters labelled the pixel as eczema (Figure 1, last column).

The segmentation performance was measured using standard metrics for computer vision classification, such as IoU, accuracy, true positive rate (TPR, aka sensitivity or recall), positive predictive value (PPV, aka precision) and F1 score, where the consensus segmentation was treated as “true” labels. We only considered skin pixels and did not exclude any images when computing the segmentation performance.

We calculated the segmentation performance for each of the four raters in turn and averaged the performance over the four raters to derive the segmentation performance of an average rater, for each image. The average rater’s segmentation performance was compared with a “naïve” segmentation performance for a “naïve” rater who would predict all skin regions to be eczema. The naïve segmentation performance corresponds to the lower bound of the segmentation performance that we would expect any segmentation algorithm to achieve.

## RESULTS

### Quality of images for eczema area segmentation

The 80 images we dealt with were from different representative AD sites: 31 images were for legs, 19 for hands, 14 for arms, 12 for feet, and 4 for the head and neck area (Table 1). The random sampling of 80 images did not select any images of trunk/back because those sites were not included abundantly enough in the original study. The average of the sum of the intensity scores (scores between 0-3) for the six signs of SASSAD (erythema, exudation, excoriation, dryness, cracking, and lichenification) was 6.3 (where the maximum is 18 = 3 × 6 signs, SD = 3.2).

The four raters fully agreed on the image quality for 23 of the 80 images. Four images were assessed to have a contradicting image quality of high and low by at least two raters, and the remaining 53 images had a combination of normal/high or normal/low. Visual inspection of the four images with a contradictory image quality, together with the reasons for the low quality provided, revealed that the textures and features of the eczema area were not well preserved or easily identifiable in those images, without obvious technical issues. 43 out of the 80 images were deemed of poor quality by at least one rater. Among those 43 images, 35 were deemed out-of-focus (Figure S2), 15 overexposed, and 14 had “other reasons” (details not given).

We computed the image quality score for each image, by averaging the quality assessed by the four raters, with “low”, “normal” and “high” being coded as −1, 0 and 1, respectively. Over the 80 images, the image quality score had a mean of −0.081 (SD = 0.45) which is close to a “normal” image quality (0). We investigated whether the image quality score could be confounded by the body regions or the severity score (the sum of the intensity scores for the six disease signs) in a linear model, but no coefficients appeared as significant (Figure S3).

### Inter-rater reliability (IRR) of eczema segmentation

The pixel-level ICC demonstrated a large image-to-image variation: 56% of the images (40/71) had *ICC* < 0.5 (“poor” agreement), 11% (8/71) with *ICC* > 0.9 (“excellent” agreement), leaving the remaining 33% with a moderate level of pixel-level agreement. The average pixel-level ICC was 0.45 (*SE* = 0.04, Figure 2), corresponding to a “poor” average agreement between raters. The pixel-level Krippendorff’s alphas (KAs) were strongly correlated with the pixel-level ICC (Fig. S3, Pearson correlation of 0.879, 95% CI: [0.812, 0.923]).

**Figure 2:**
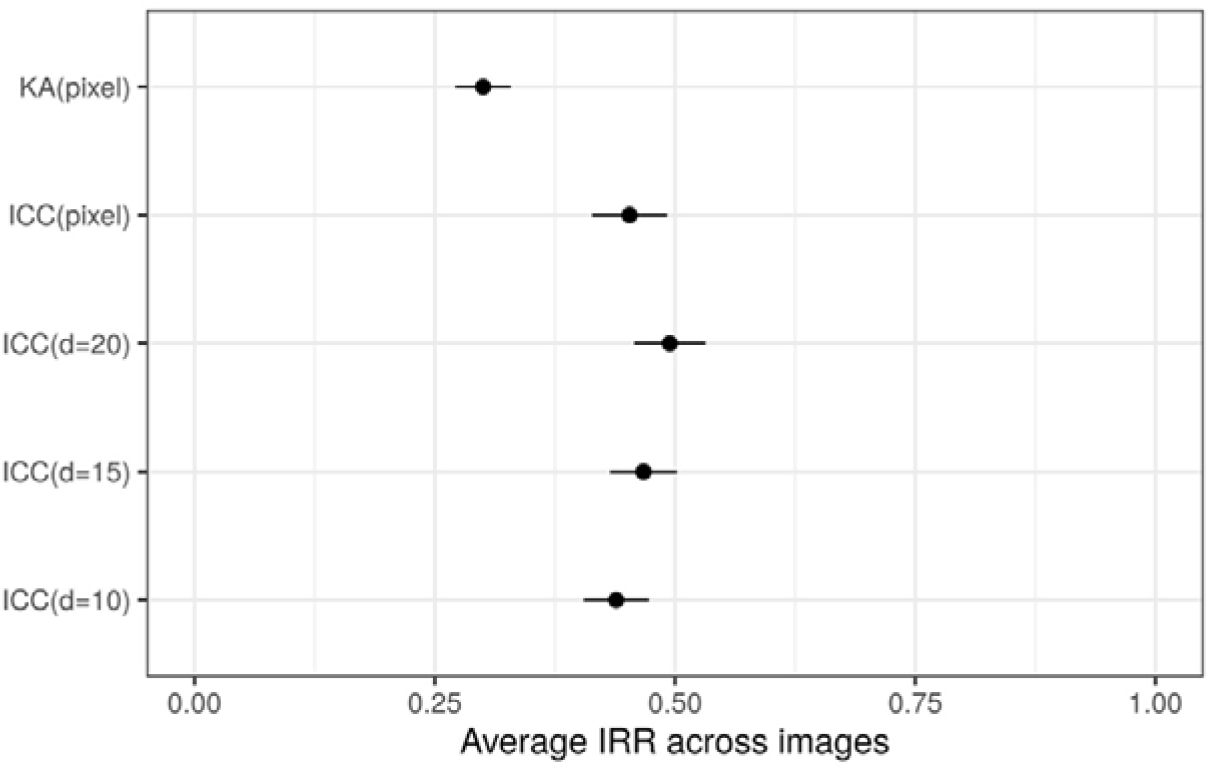
Average (± SE) pixel- and area-level IRR estimates (the higher the better). KA(pixel) and ICC(pixel) correspond to pixel-level IRR metrics, and other ICC corresponds to area-level IRR metrics, where d describes the image resolution. A larger d corresponds to smaller area of interest.

The area-level ICC for different resolutions of the images were strongly correlated with each other and with pixel-level ICC (Figure S4). The average area-level ICC was not significantly different from the average pixel-level ICC and corresponded to a “poor” average agreement between raters (Figure 2). The extent ICC was 0.440 (95% CI: [0.313, 0.555]) confirming that the raters could not agree on the proportion of eczema in the images, even without considering the eczema segmentation.

We investigated whether the IRR was confounded by body regions, severity scores, average labelling time by the four raters, and image quality scores, using a linear model with IRR metrics as the dependent variables (Figure S5). The only significant effect was found for the image quality scores, with a *higher* quality associated with a *lower* ICC. This counter-intuitive result may be explained by the fact that the raters did not attempt a precise segmentation on lower quality images resulting in a higher agreement.

Our sensitivity analysis did not show significant differences in pixel-level ICC when one of the raters was removed in turn from the analysis (Figure S6). Our results are not driven by the segmentation labels of a particular rater, including the most experienced rater (rater 1).

### Performance for eczema area segmentation

We estimated the average rater’s segmentation performance and compared it with a “naïve” segmentation performance achieved by a “naïve” rater who segments all skin regions as eczema (Figure 3). The performance of the average rater’s segmentation was always better than that of a naïve segmentation, except for the true positive rate metric (TPR) which does not penalise false positives that the naïve segmentation produces by many. None of the metrics for the average rater’s performance was close to a perfect score of 1, which would have been expected if the IRR were excellent and any raters’ segmentation could be used interchangeably. Our estimation suggested that an average rater can segment eczema with an accuracy of 80.6 ±1.5 % (averaged across images) compared to the consensus of their peers.

**Figure 3:**
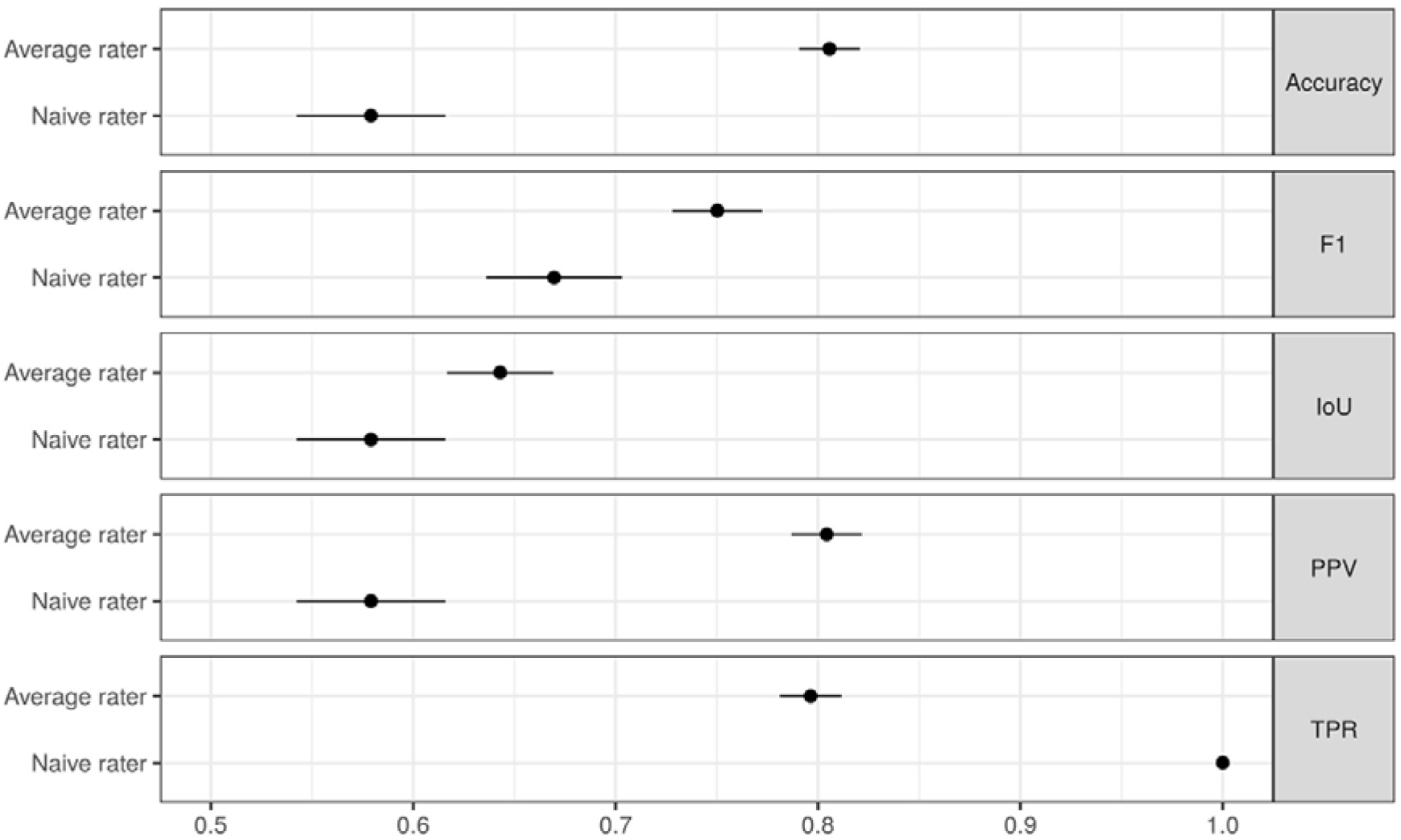
The segmentation performance of the average rater and the naïve rater (mean ± *SE* across images, the higher the better). IoU: intersection over union; PPV: positive predicted value (precision); TPR: true positive rate (sensitivity, recall).

The difference between the naïve and the average raters’ performance varies across metrics, with a greater difference for Accuracy, PPV and TPR than for the F1 score and IoU (Figure 3). These results suggest that not all metrics may be appropriate to monitor improvement in the segmentation performance of a ML algorithm and discourage the use of F1 score and IoU, as they demonstrated a smaller difference between the naïve and the average raters’ performance. A narrow difference makes it difficult to detect the improvement in the segmentation performance of a ML algorithm that is likely to be above the naïve performance and below the average rater’s performance.

## DISCUSSION

In this study, we investigated the inter-rater reliability (IRR) of eczema segmentation from digital images. Four dermatologists (raters) segmented eczema lesions in 80 images collected in a previously published clinical trial (Figure 1). The IRR of eczema segmentation varied from image to image with a poor agreement between the raters on average (Figure 2). We also estimated the segmentation performance for an average rater and a naïve rater who segments all skin regions as eczema (Figure 3). Those segmentation performances could be used to benchmark eczema segmentation algorithms and to choose the most appropriate metric to monitor the performance of eczema segmentation algorithms.

Our results highlight the difficulty of detecting eczema lesions from digital images consistently, raising questions on the validity of machine learning (ML) models to automatically assess eczema severity if they rely on eczema area segmentation without accounting for raters’ potential biases. The results are perhaps not surprising given that AD is “ill-defined erythema with surface change” by definition [15] and that clinical assessment of affected areas is challenging [8].

The problem of the poor IRR in the segmentation data for training ML models could be addressed in several ways. For example, we can design end-to-end ML models for automatic assessment of eczema severity without relying on eczema segmentation masks provided by dermatologists, and instead work with original images or images with background removed (using skin segmentation masks) and let the algorithm identify eczema regions by itself. Other possibilities include using eczema segmentation algorithms that can be trained on noisy segmentation labels [7], or trying to improve eczema segmentation labels. Improving the quality of eczema segmentation could be achieved by better training of raters, such as providing feedback on a reference set of training images until a certain level of ICC is achieved. Averaging the segmentation from multiple independent raters may also help. For example, the ICC of an average (consensus) segmentation by nine independent raters will be 0.9 (excellent) if we assume the ICC of an individual segmentation is 0.5 (poor) [13]. It may also be possible to ask fewer raters or even non-experts to segment eczema and take advantage of the raters’ systematic biases using crowdsourcing models such as a Dawid-Skene model.

There were several limitations in this study. Firstly, our randomly selected 80 images from a published clinical trial mostly contained images of white skin tones. Further research is needed to investigate the quality of eczema segmentation for darker skin tones. For example, erythema is more likely to appear violaceous or just dark brown in darker skin tones, making the delineating of the inflamed border potentially more challenging even if the lesion is not completely missed [16]. Collecting images of eczema on darker skin tones is also relevant for developing ML algorithms that tend to be more accurate on the skin types they were trained on [17]. Secondly, the IRR metrics used in this study did not consider the spatial structure of the image, while neighbouring pixel labels are unlikely to be independent. This could be addressed with additional pre-processing such as computing local extent values using a kernel smoother (e.g. Gaussian blurring), which would also avoid “compressing” the masks when computing the area-level IRR. Our results were nonetheless consistent between pixel-level, area-level and extent IRRs, highlighting the robustness of our conclusions. Finally, it would be valuable to estimate the intra-rater reliability of AD segmentation in digital images, i.e. to what extent raters identify the same lesions on different occasions consistently. Poor intra-rater reliability may also be detrimental to the development of machine learning models relying on AD segmentation data.

In conclusion, this study demonstrated that the AD segmentation in digital images is highly rater-dependent even between dermatologists. Such limitations need to be taken into consideration when the AD segmentation data are used to train machine learning algorithms that assess eczema severity.

## Data Availability

All the codes are available at https://github.com/ghurault/IRR-eczema-images.

## DATA AVAILABILITY

All the codes are available at https://github.com/ghurault/IRR-eczema-images.

## ACKNOWLEDGMENTS

We thank Prof Kim S Thomas and the SWET study team for sharing the SWET dataset. The SWET trial was funded by the NIHR Health Technology Assessment Programme.

