## Supplementary Figures for "Detecting eczema areas in digital images: an impossible task?"

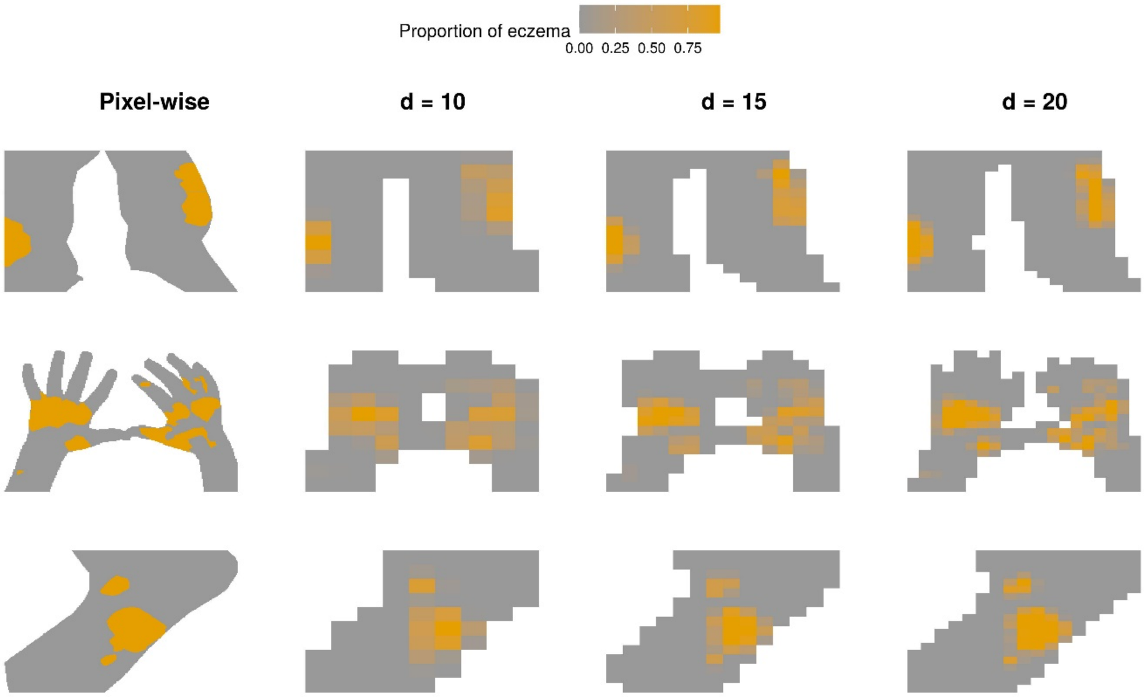


**Figure S1**: Illustration of area-level segmentation compared to pixel-level segmentation, for the same images as in Figure 1, for the segmentation for rater 1. Rows correspond to images and columns to pixel- and area-level segmentation for different image resolutions ($d$).

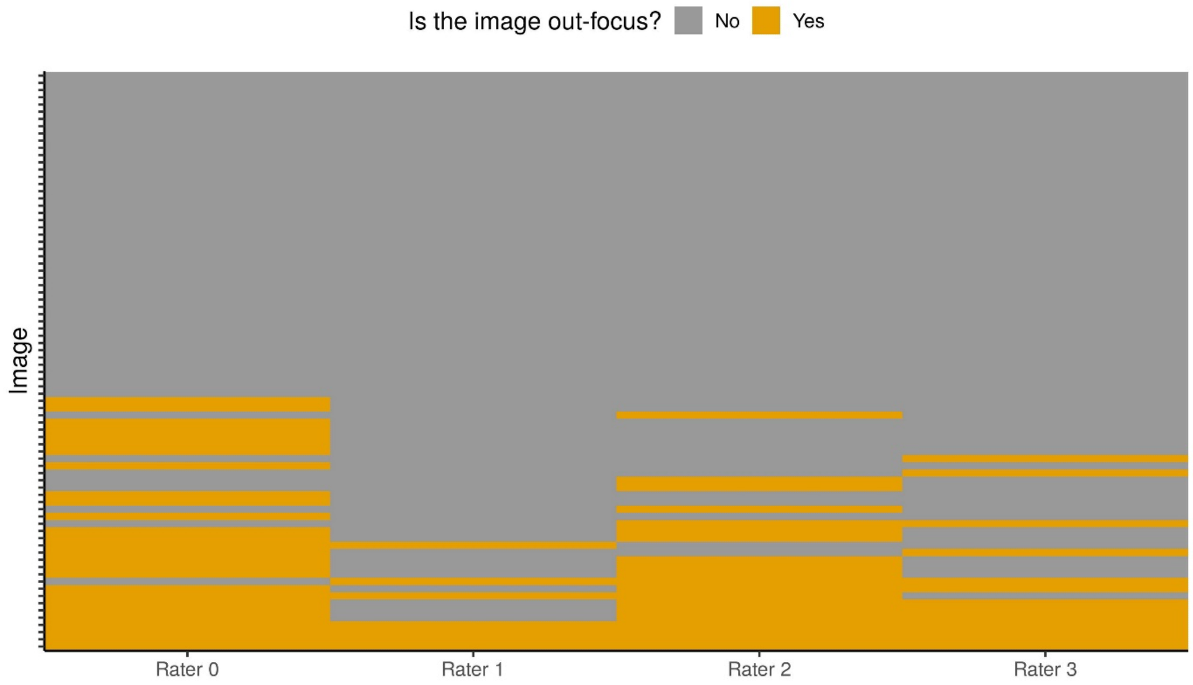


**Figure S2**: Distribution of out-of-focus assessments by the four raters (x-axis) for each image (y-axis). The orange represents that the rater deemed the image out-of-focus. 17 images were deemed out-of-focus by only one rater, 8 images by two raters, 6 images by three raters and 4 images by all the four raters.

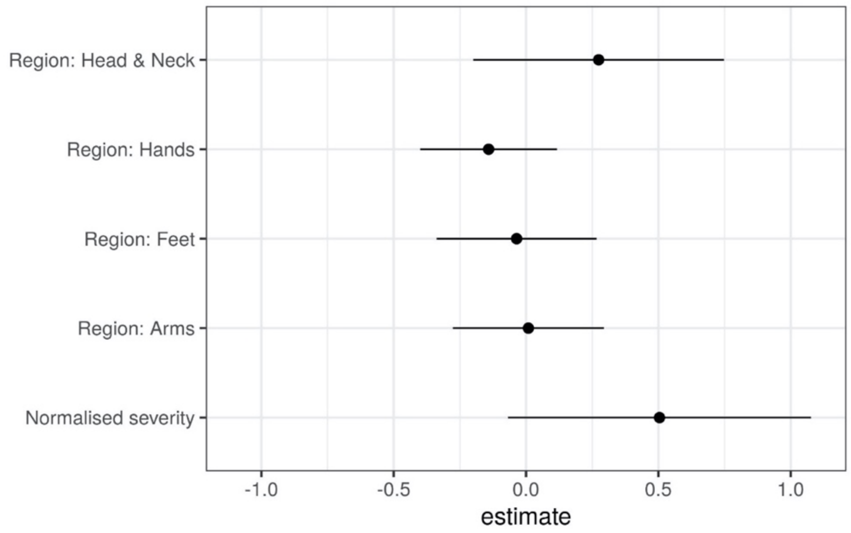


**Figure S3:** Estimated coefficients (and 95% CI) for variables in a linear model that predicts the mean image quality score across raters. The coefficients for the regions quantify the difference in the intercept from that of the default region (legs).

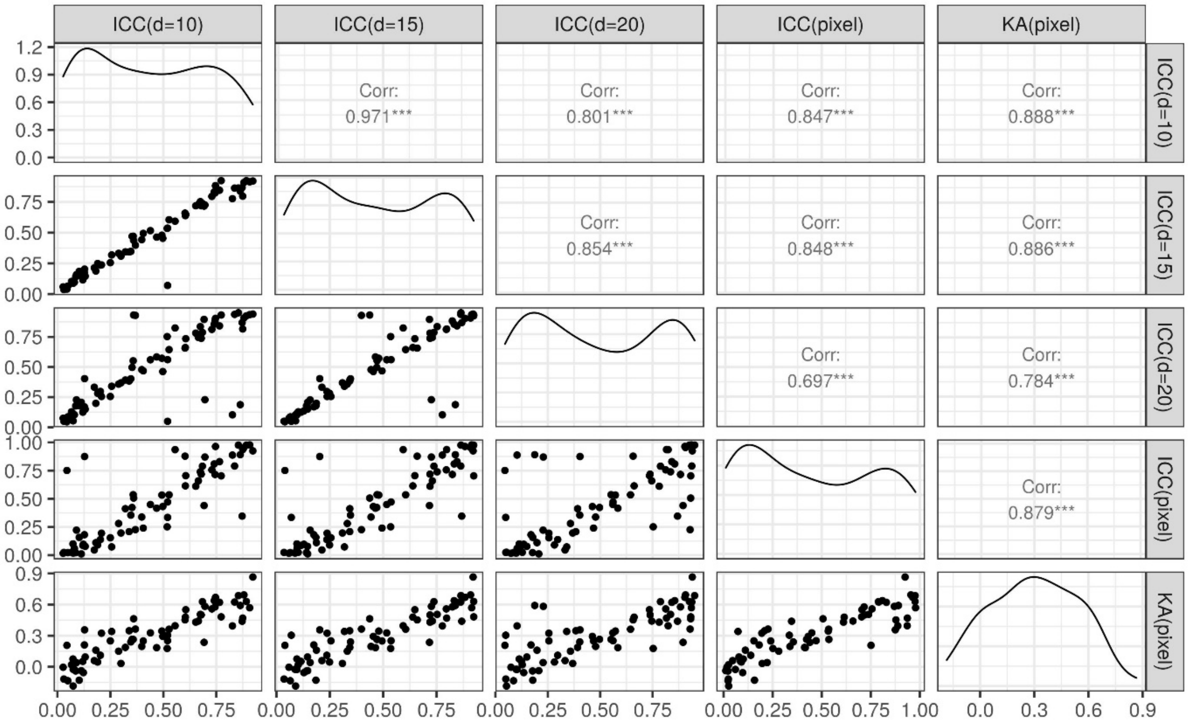


**Figure S4:** Comparison between the IRR metrics considered in this study (scatter plots, density plots and Pearson correlations).

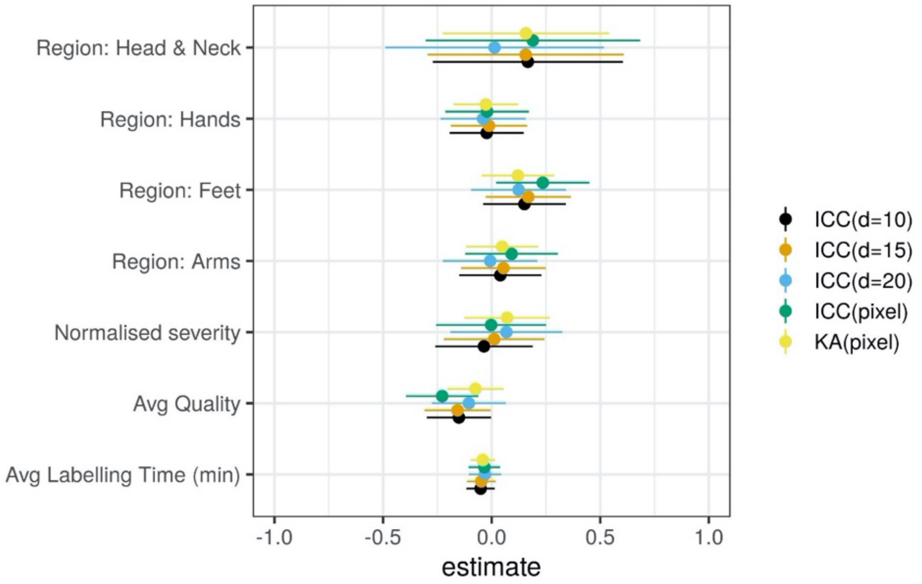


**Figure S5:** Estimated coefficients (and 95% CI) for the variables in a linear model that predicts IRR metrics. The variables were normalised to a sensible scale for a fair interpretation of the effect sizes. The coefficients for the regions quantify the difference in intercept from that of the default region (legs).

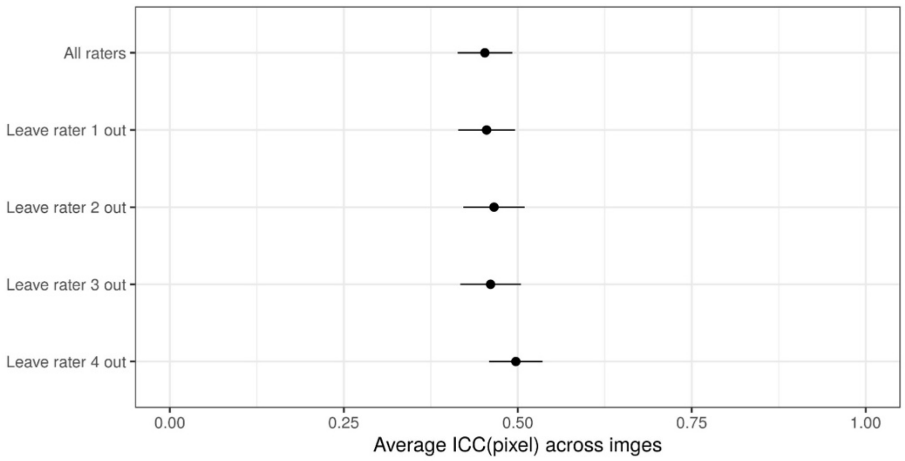


**Figure S6:** Leave-one-rater out sensitivity analysis of the average pixel-level ICC measure.
